# Using routinely collected hospital data to investigate healthcare worker mobility and patient contacts within a UK hospital during the COVID-19 pandemic

**DOI:** 10.1101/2022.06.10.22276247

**Authors:** Jared K Wilson-Aggarwal, Nick Gotts, Wai Keong Wong, Chris Liddington, Simon Knight, Moira J Spyer, Catherine Houlihan, Eleni Nastouli, Ed Manley

## Abstract

Movement and contacts are central to the transmission of infectious diseases and, within the hospital setting, healthcare worker (HCW) mobility and their contact with patients play an important role in the spread of nosocomial disease. Yet data relating to HCW behaviours associated with mobility and contacts in the healthcare environment are often limited. This paper proposes a framework for integrating several electronic data sources routinely-collected by modern hospitals, to enable the measurement of HCW behaviours relevant to the transmission of infections. Using data from a London teaching hospital during the COVID-19 pandemic, we demonstrate how, at an aggregate level, electronic medical records (EMRs) and door access logs can be used to establish changes in HCW mobility and patient contacts. In addition, to show the utility of these data sources in supporting infection prevention and control (IPC), we investigate changes in the indirect connectivity of patients (resulting from shared contacts with HCWs) and spatial connectivity of floors (owing to the movements of HCWs). Average daily rates of patient contacts are computed and found to be higher throughout the pandemic compared to that pre-pandemic, while the average daily rates of HCW mobility remained stable until the second wave, where they surpassed pre-pandemic levels. The response of HCW behaviour to the pandemic was not equal between floors, whereby the highest increases in patient contacts and mobility were on floors handling the majority of COVID-19 patients. The first wave of COVID-19 patients resulted in changes to the flow of HCWs between floors, but the interconnectivity between COVID-19 and non COVID-19 wards was evident throughout the pandemic. Daily rates of indirect contact between patients provided evidence for reactive staff cohorting, whereby indirect contact rates between COVID-19 positive and negative patients were lowest during peaks in COVID-19 hospital admissions. We propose that IPC practitioners use these routinely collected data on HCW behaviour to support infection control activities and to help better protect hospital staff and patients from nosocomial outbreaks of communicable diseases.

## Introduction

Human mobility and contact are significant drivers for the transmission of communicable diseases, such as severe acute respiratory syndrome coronavirus 2 (SARS-CoV-2) that resulted in the COVID-19 pandemic (Buckee et al., 2021). While passively collected mobile phone and app-derived GPS trajectory data provide an indication of populations’ mobility and social mixing patterns (Ross et al., 2021), only broad regional generalisations can be drawn. Transmission of SARS-CoV-2 occurs through close proximity between infectious and susceptible individuals due to either direct contact or respiratory aerosols in the shared space (Rahman et al., 2020). Therefore, insights into behaviours at a fine scale, such as within indoor environments, are also required to deepen our understanding of behaviours associated with the transmission of infections, and improve our ability to identify and prevent transmission events. This is particularly relevant for healthcare settings, where infection outbreaks present a significant risk to vulnerable patients through increased morbidity and mortality.

The concern in relation to infection transmission within hospital environments extends more widely than COVID-19, and includes other healthcare-associated infections (HAIs). The impact of HAIs on healthcare systems is considerable, resulting in staff illness, complications in patient outcomes and increasing healthcare costs. In England between 2016-2017, HAIs were estimated to have caused >28,000 deaths, contributed to 21% of hospital bed days, resulted in >79,000 days of absence in frontline HCWs and cost the NHS an estimated £2.7 billion (Guest et al., 2020). The surveillance prevention and control of HAIs is a challenge as granular data are often limited and the transmission pathways are highly variable; dependent on the epidemiology of the pathogen, be it bacterium, virus or fungus (Khan et al., 2015, 2017).

Nosocomial infections in patients are well defined, as are frameworks for their prevention and control (Loveday et al., 2014). To manage HAIs in patients, practitioners responsible for infection prevention and control (IPC) frequently use passive data sources that are routinely collected, such as medical records. These data sources provide information on the patient’s location within the hospital and their contacts with staff, which can be used to support surveillance, mapping patient trajectories and contact tracing (Murray et al., 2017; Price et al., 2021; Rewley et al., 2020).

Historically these data sources have been handled manually, using time intensive frameworks that prevent their use in real-time. Hospitals that have moved to digital systems have seen an increase in the effectiveness and efficiency of patient focused IPC, through improved availability of data resources and reduced burdens of manual data collection and processing (Chen et al., 2019; Russo et al., 2018). However, while these data streams are well established for patient focused activities, those for the management of HAIs in HCWs are relatively underdeveloped. This is surprising given that, like patients, HCWs are at risk of both acquiring and facilitating the transmission of HAIs (Huttunen & Syrjänen, 2014).

The rapid spread of SARS-CoV-2 has emphasized the need to protect front-line HCWs. Early in the pandemic the prevalence of COVID-19 infection for HCWs was high, with one London hospital reporting infection in 44% of HCWs (Houlihan et al., 2020) and a global estimation of 11% of HCWs infected with the virus (Gómez-Ochoa et al., 2021). What’s more, the risk of infection for HCWs varied between roles and spatially, with a higher risk of infection for those working in non-emergency wards and for nurses (Gómez-Ochoa et al., 2021). Nosocomial outbreaks of SARS-CoV-2 result from a small number of highly infectious individuals, and transmission chains may include HCWs among the likely ‘super-spreaders’ (Lumley et al., 2021). Behavioural processes, such as contact and mobility patterns, generate heterogeneity in the transmission of communicable diseases (Arthur et al,. 2017; Buckee et al., 2021) and, similar to the management of HAIs in patients, passively collected data on the within-hospital behaviours of HCWs can contribute to a more informed and rapid response to outbreaks.

HCW behaviours have been investigated using surveys (English et al., 2018), observations (Weigl et al., 2009; Westbrook et al., 2011) and tracking technologies (Butler et al., 2018; Hertzberg et al., 2017; Oussaid et al., 2016; Vanhems et al., 2013), but these data collection methods are often prohibitively time intensive, expensive, or only provide a snapshot view that is not hospital wide. Electronic medical records (EMRs) have also been previously used to investigate HCW space use and patient contacts (Curtis et al., 2013; Illingworth et al., 2022), but they are either optimised for reconstructing patient trajectories or suffer from high spatial uncertainty. Additional databases, such as door access logs could complement EMRs by supplementing spatiotemporal information on HCW mobility. These data sources are analogous in nature to the passively-collected spatial data from mobile phone records, which were used during the COVID-19 pandemic to demonstrate the effectiveness of movement restrictions in reducing contact rates, and subsequently lowering levels of community transmission (Nouvellet et al., 2021). Using the routinely collected hospital data as an indicator for HCW behaviour provides opportunities to enhance evidence based IPC in a similar way; supporting contact tracing efforts, validating transmission pathways and helping to monitor the effectiveness of interventions in the hospital.

As with other communicable diseases, IPC interventions to prevent nosocomial outbreaks of COVID-19 include hand washing, the use of personal protective equipment (PPE), limiting the traffic of people in the hospital, and cohorting staff and patients (Ahmad & Osei, 2021). The routinely collected data cannot identify or monitor all HCW behaviours that are epidemiologically relevant, but can indicate their level of space use within the hospital, their frequency of movement, the number of patients they contact and the frequency of patient contact. As behavioural markers these metrics provide quantitative measures for IPC interventions aimed specifically at reducing the spatial connectivity of spaces (e.g. by restricting staff access/flow to areas) and social connectivity of individuals in the hospital (e.g. through patient and/or staff cohorting). The data can therefore be used to assess the extent to which interventions targeted towards HCW mobility and patient contacts have been successful in achieving their aim, or in determining opportunities for improvement.

This paper outlines a framework for the use of routinely-collected hospital data in the measurement of HCW behaviour. We describe (1) the integration of diverse digital data sources for the quantification of HCW mobility and patient contact within the hospital setting, and (2) demonstrate the use of these data sources in supporting IPC activities through a series of analyses. Specifically, we use data from a London Hospital during the COVID-19 pandemic to measure at an aggregate level how (i) the temporal and spatial patterns of HCW mobility and patient contacts, (ii) spatial connectivity (flow between floors) and (iii) indirect contacts between patients (through shared HCW contacts) changed during the COVID-19 pandemic.

## Methodology

### Study site and context

University College London Hospital NHS Trust (UCLH) is a large acute and tertiary referral academic hospital located in central London. The Main UCLH building is comprised of a central Tower that has 19 floors (floors -2 to 16) and is linked to two other buildings; the Podium and the Elizabeth Garett Anderson (EGA) Wing.

In this analysis, we only considered data for the Tower building at UCLH. Here we describe floors within the Tower by the ward/department that predominantly occupies it; the basement (floor-2), imaging (floor -1), emergency department (ED on floor 0), acute medicine unit (AMU on floor 1), day surgery (floor 2), critical care (floor 3), plant (floor 4), nuclear medicine (floor 5), short stay surgery (floors 6), hyper-acute stroke unit (HASU on floor 7), respiratory & infectious diseases (floor 8), general surgery (floor 9), care of the elderly (CoE on floor 10), paediatrics (floor 11), adolescents (floor 12), oncology (floor 13), head and neck (floor 14), private wards (floor 15) and haematology (floor 16).

During the pandemic the UCLH Tower became a key site for COVID-19 care in London, and the peaks in the number of COVID-19 patients in the Tower (Figure 1C), closely followed that reported for all London hospitals (as downloaded from gov.uk; r = 0.97). To investigate changes in the daily number of events in different stages of the pandemic, we manually identified four distinct time periods based on the number of COVID-19 patients in the Tower. The first time period identified was January 1^st^– February 28^th^ 2020, which was considered pre-pandemic or the ‘baseline’, as this was prior to the substantial rise in COVID-19 admissions in the hospital, so ‘normal’ patterns of movement and patient contacts would be expected. The second time period was March 1^st^– June 30^th^ 2020, when the ‘first wave’ of COVID-19 hospital admissions was experienced, during which the WHO declared a pandemic (March 11^th^ 2020) and the first national lockdown in England was announced (23^rd^ March 2020). The third time period was July 1^st^– August 31^st^ 2020, which represented the ‘summer lull’, where the number of COVID-19 patients in London hospitals remained at a low level and community interventions were eased. The fourth time period was November 1^st^ 2020 - March 31^st^ 2021, when the ‘second wave’ of COVID-19 hospital admissions occurred, the second national lockdown was announced (5^th^ November 2020) and the mass-vaccination programme began (December 8^th^ 2020).

**Figure 1:**
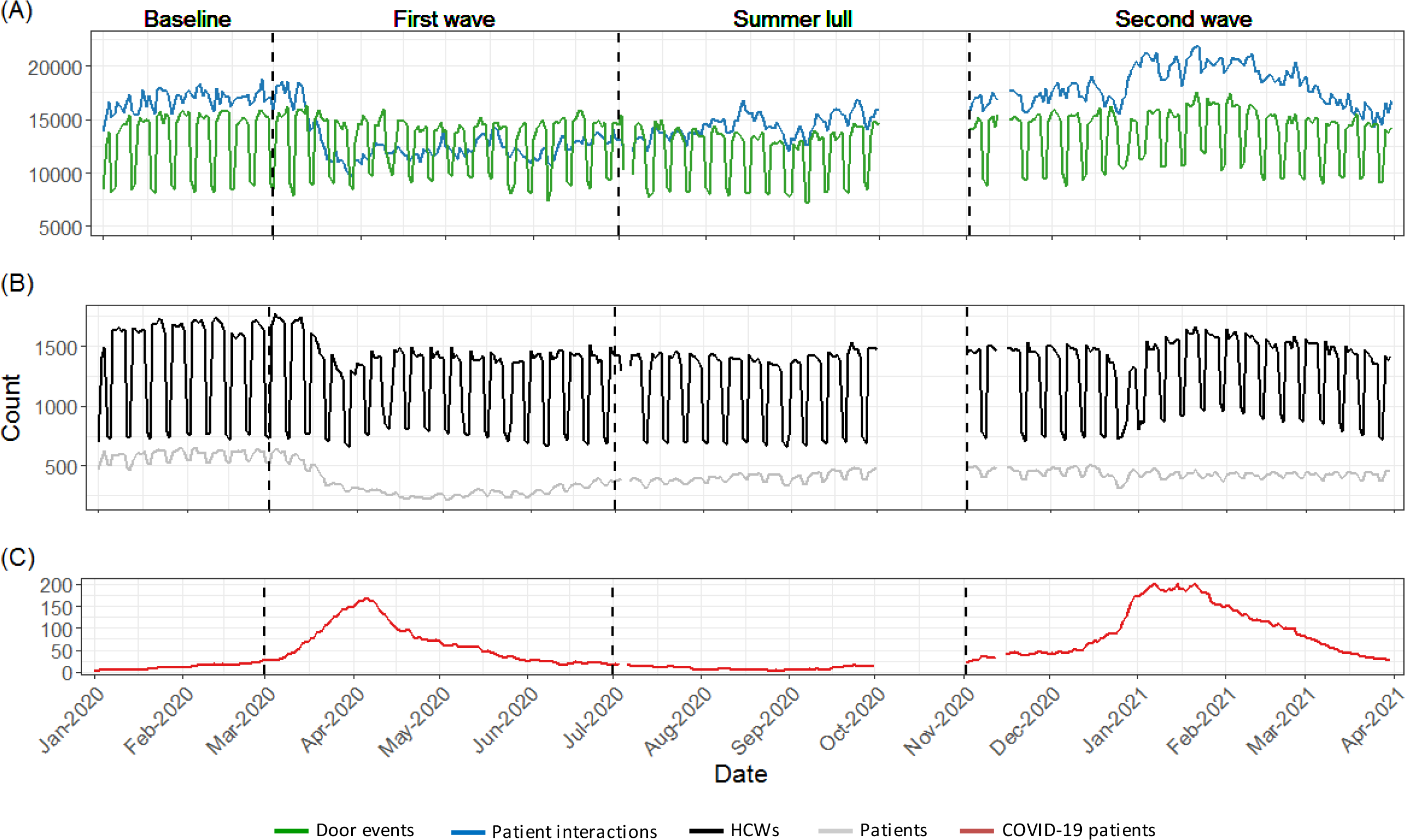
Daily counts for indicators of healthcare worker behaviour, staffing levels and patient numbers in the Tower building of University College London hospital during the COVID-19 pandemic. Plot A shows the activity of healthcare workers in the hospital as characterised by the daily count of door events (green) and patient contacts (blue) logged in routinely collected electronic data sources. Plot B shows daily counts for the number of healthcare workers (black) and patients (grey) identified in the data. Plot C shows the number of COVID-19 patients in the hospital, which was used to determine the different stages of the pandemic; Baseline (pre-pandemic), first wave, summer lull and second wave. Data for October 2020 was not available.

### Data sources

The data sources used in this study were selected on the basis of providing spatial and temporal indicators of staff movement and patient contacts within the hospital. For clarity, Table 1 provides definitions of terms relating to behavioural processes that are investigated in this study.

**Table 1.** Definitions and data sources for behavioural processes relating to healthcare worker activity in the healthcare environment.

| Behavioural process | Definition | Data source |
| --- | --- | --- |
| Mobility | The frequency of movement exhibited by individuals/populations between discrete locations. | Security door access logs |
| Direct patient contact | Face to face interactions between healthcare workers and patients. | Electronic medical records |
| Indirect contact | The secondary contact between individuals resulting from their direct contact with others. | Electronic medical records |
| Social connectivity | The relationship between individuals, as determined by their direct and/or indirect contacts. | Electronic medical records |
| Spatial connectivity | The relationship between discrete locations, as determined by the spatial activity of individuals. | Security door access logs & electronic medical records |

#### Electronic medical records (EMRs)

Patient contact events in EMRs were extracted from Epic, a privately owned hospital system used by UCLH for managing medical records. While Epic contains a large volume of data on patient diagnosis and treatment, we only use specific fields that provide information on the spatial and temporal attributes of within-hospital contacts between staff and patients. Data fields included the datetime of events, a description of the location (bed ID and floor), an indicator of the event type, anonymous identifiers for the patient, pseudonymous identifiers for the HCW and the COVID-19 status of the patient at the time of the event (0/1).

#### Card access logs

Door events were extracted from the database for security door access logs, known in this context as CCure. Data fields included the datetime of events, a description of the location (door ID and floor), direction of passage (in/out), status (accepted/rejected) and a pseudonymous staff identifier.

### Data Processing

#### Data cleaning

All data processing was conducted in R (R Core Team, 2020). Data for events outside the UCLH Tower were discarded. Data for the month of October 2020, a weekend in July and another in November were also discarded, as records either could not be extracted or had an unusually low number of events (indicating an issue with extraction). Contact events in EMRs that did not require face to face contacts (e.g. ‘Telephone’ or ‘Letter’) were excluded. Door events with a rejected status were removed along with duplicate events in the same direction that were within 60 seconds of each other. Two types of lift (or elevator) events were present in the door access logs; ‘Lift Calls’ where a card is used to request a lift, and ‘Lift commands’ where a card is used before selecting which floor to go to. All ‘Lift Call’ events were removed as they overinflate the number of movement events for individuals using lifts (because some individuals may make multiple and repeated lift calls while waiting for a lift).

#### Aggregate measures

Staffing levels, |*H*|, were determined by summing the number of distinct HCWs, *h*, in the set of HCW IDs identifiable in the routinely collected data, *H*. Staffing levels were calculated for each day, *t*, stage of the pandemic, *Stage*, and for each floor, *f*, and the entire building. For each stage of the pandemic the mean daily staffing levels, |*H̅*|, were calculated for the entire building and each floor. Similarly, patient levels, |*P*|, were calculated by summing the number of distinct patients, *p*, in the set of patient IDs identifiable in the data, *P*, for each day, and the mean daily patient levels, *P̅*, calculated for the entire building, and for each floor, during each stage of the pandemic. The same metrics were also calculated using the subset of patients known to be positive for COVID-19, *P*^*Positive*^.

Door events, *m*, were used as an indicator of HCW behaviour in terms of their mobility, where *M* is the full set of door events. The number of door events, |*M*|, was used as an absolute measure of HCW mobility and was calculated for the entire building and each floor on each day. There was a strong correlation between daily HCW mobility and daily staffing levels (r = 0.94) and therefore, to control for changes in staffing levels, the rate of mobility, *Mr*, was calculated as a function of staffing levels, where:

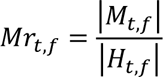

To compare mobility levels between stages of the pandemic, the mean daily mobility, *M*, and the mean daily rate of mobility, *Mr̅*, were calculated for the entire building and for each floor during the different stages of the pandemic.

Patient contact events, *c*, were used as an indicator of HCW behaviour in terms of patient engagement where *C* is the full set of patient contacts. The number of patient contacts, |*C*|, was used as an absolute measure and was calculated for the entire building and each floor on each day. There was a correlation between daily patient contacts and daily patient levels (r = 0.65), and therefore we also calculated the daily rate of patient contacts, *Cr*, as a function of patient levels, where:

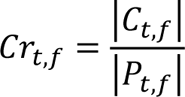

To compare levels of patient engagement between stages of the pandemic, the mean daily number of patient contacts, *C̅*, and the mean daily rate of patient contacts, *Cr*, were calculated for the entire building and for each floor during the different stages of the pandemic.

To investigate the weekly and hourly patterns of mobility and patient engagement, a count for the number of door events and patient contacts was made for each hour, *hr*, of each weekday, *w*, and separately for the different stages of the pandemic. These counts were then weighted by dividing them by the number of days each weekday appeared in the dataset.

#### Changes in time and space

To investigate how the measures of daily HCW behaviour, staffing levels and patient levels differed from that pre-pandemic (baseline), on each floor and within the entire building, the normalised difference to baseline, *N*, was calculated for each day e.g. normalised difference to baseline for HCW mobility across the entire building, *N*_*t*_^*M*^, where:

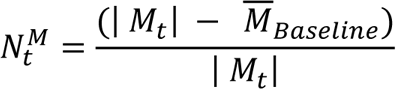

The normalised difference to baseline was also calculated for the averaged values for HCW behaviour during each stage of the pandemic. *N* can be interpreted as proportional change, but is presented as percentage change in the results.

#### Spatial connectivity

To assess the relationship between floors, a dyadic analysis was conducted for the different stages of the pandemic. For each spatial dyad (e.g. floors 1 & 2, floors 1 & 3 etc.) and using both door events and patient contacts, the number of HCWs that were active on both floors in any single day was extracted where:

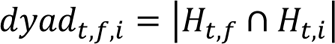

The index of the second floor in the dyad is denoted *i*. The resulting matrix was then treated as a weighted network with the diagonal set to zero. The Louvain clustering algorithm was used to identify floors that had stronger links. Louvain clustering is a hierarchical greedy modularity maximization-based approach (Blondel et al., 2008) and was implemented using the R package ‘igraph’ v1.2.7 (Csardi & Nepusz, 2006). Lift events were excluded from this analysis as it was not possible to identify the floor on which they occurred.

#### Patient connectivity

Patient connectivity, *S*, was determined by identifying the number of COVID-19 negative patients each patient was indirectly in contact with through shared contacts with the same HCWs on the same day. This was achieved by first identifying the set of HCWs that had contact with the *j*th patient on each day, where:

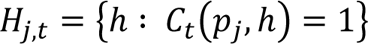

Next the set of patients not known to be positive for COVID-19, *P*^*Negative*^, and that had also been in contact with any of the HCWs in *H*_*j,t*_ were identified, where:

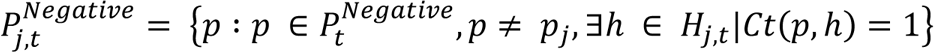

*S* was then calculated for each patient as a proportion of all patients not known to be positive for COVID-19, and expressed as a percentage in the results where:

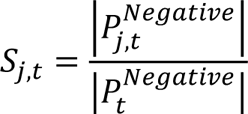

For each day and stage of the pandemic, we made separate calculations for the average patient connectivity of patients not known to be positive for COVID-19, *S̅^Negative^*, and COVID-19 positive patients, *S̅^Positive^*.

## Results

Data were analysed for 7,975 HCWs that had logged door events and/or patient contacts in the UCLH Tower building between January 2020 and March 2021. In total, 5,510,359 door events and 6,574,590 patient contacts were recorded. During the entire observation period, 21,133 patients were detected in the routinely collected data, of which 8% were positive for COVID-19. Table 2 provides a summary for the different stages of the pandemic.

**Table 2:** Summary for indicators of healthcare worker staffing levels, mobility and patient contacts in the Tower building of University College London Hospital during the COVID-19 pandemic. For each stage of the pandemic, the duration in days is reported along with the total number and average daily count of healthcare workers, patients, patients positive for COVID-19 (COVID-19^+^), door events, patient contacts, and contacts with COVID-19^+^. The mean daily rate of door events (per healthcare worker) and mean daily rate of patient contacts (per patient) are also reported. For counts involving COVID-19^+^, the percentage of all patients/contacts are provided in brackets.

|  | Baseline<br>Jan – Feb<br>2020 | First wave<br>March – June<br>2020 | Summer lull<br>July – Aug<br>2020 | Second wave<br>Nov 2020 –<br>March 2021 | Overall |
| --- | --- | --- | --- | --- | --- |
| Days | 61 | 123 | 91 | 147 | 422 |
| No. of staff $\vert H\vert $ | 4,763 | 5,526 | 5,089 | 6,107 | 7,975 |
| Avg. daily staffing levels $\bar{H}$ | 1,373 | 1,235 | 1,213 | 1,302 | - |
| No. of patients $\vert P\vert $ | 6,522 | 5,695 | 5,814 | 7,926 | 21,133 |
| No. of COVID patients $\vert P^{Positive}\vert $ (% of all patients) | 35 (1%) | 565 (10%) | 63 (1%) | 1082 (14%) | 1,688 (8%) |
| Avg. daily patient levels $\bar{P}$ | 587 | 330 | 403 | 434 | - |
| No. of events $\vert M\vert $ | 795,057 | 1,598,378 | 1,095,257 | 2,021,667 | 5,510,359 |
| Avg. daily no. of events $\bar{M}$ | 13,034 | 12,995 | 12,036 | 13,753 | - |
| Avg. daily rate of events $\overline{Mr}$ | 10 | 11 | 10 | 11 | - |
| No. of contacts $\vert C\vert $ | 1,025,271 | 1,572,340 | 1,305,838 | 2,671,141 | 6,574,590 |
| Avg. daily no. of contacts $\bar{C}$ | 16,808 | 12,783 | 14,350 | 18,171 | - |
| Avg. daily rate of contact $\overline{Cr}$ | 29 | 41 | 36 | 42 | - |
| No. of contacts with COVID-19+ $\vert C^{Positive}\vert $ (% of all contacts) | 37,393 (4%) | 527,986 (34%) | 42,801 (3%) | 996,440 (37%) | 1,604,620 (24%) |
| Avg. daily no. of COVID-19+ contacts $\bar{C}^{Positive}$ | 613 | 4293 | 470 | 6779 | - |
| Avg. daily rate of COVID-19+ contacts $\overline{Cr}^{Positive}$ | 51 | 69 | 50 | 73 | - |

In the following sections we describe the temporal and spatial patterns in the behaviour of HCWs, and how these changed throughout the pandemic. We also describe epidemiologically relevant changes in the patterns of spatial connectivity and indirect contacts between patients.

### Temporal dynamics

During the baseline period, the daily number of door events and patient contacts showed clear temporal regularity, whereby the number of events was highest during weekdays (Figure 1A). These peaks were in line with the daily pattern of staff and patient numbers in the hospital, both of which also showed weekday highs (Figure 1B). The hourly number of door events were highest on weekdays between 7am-5pm, but this peak was less prominent at weekends (Figure 2A). Regardless of the day, the hourly number of patient contacts peaked once at 10am and again at 6pm (Figure 2E). These temporal patterns demonstrate the utility of the routinely collected data in depicting the global activity levels of HCWs, which will underline the nature of staff working patterns within the hospital.

**Figure 2:**
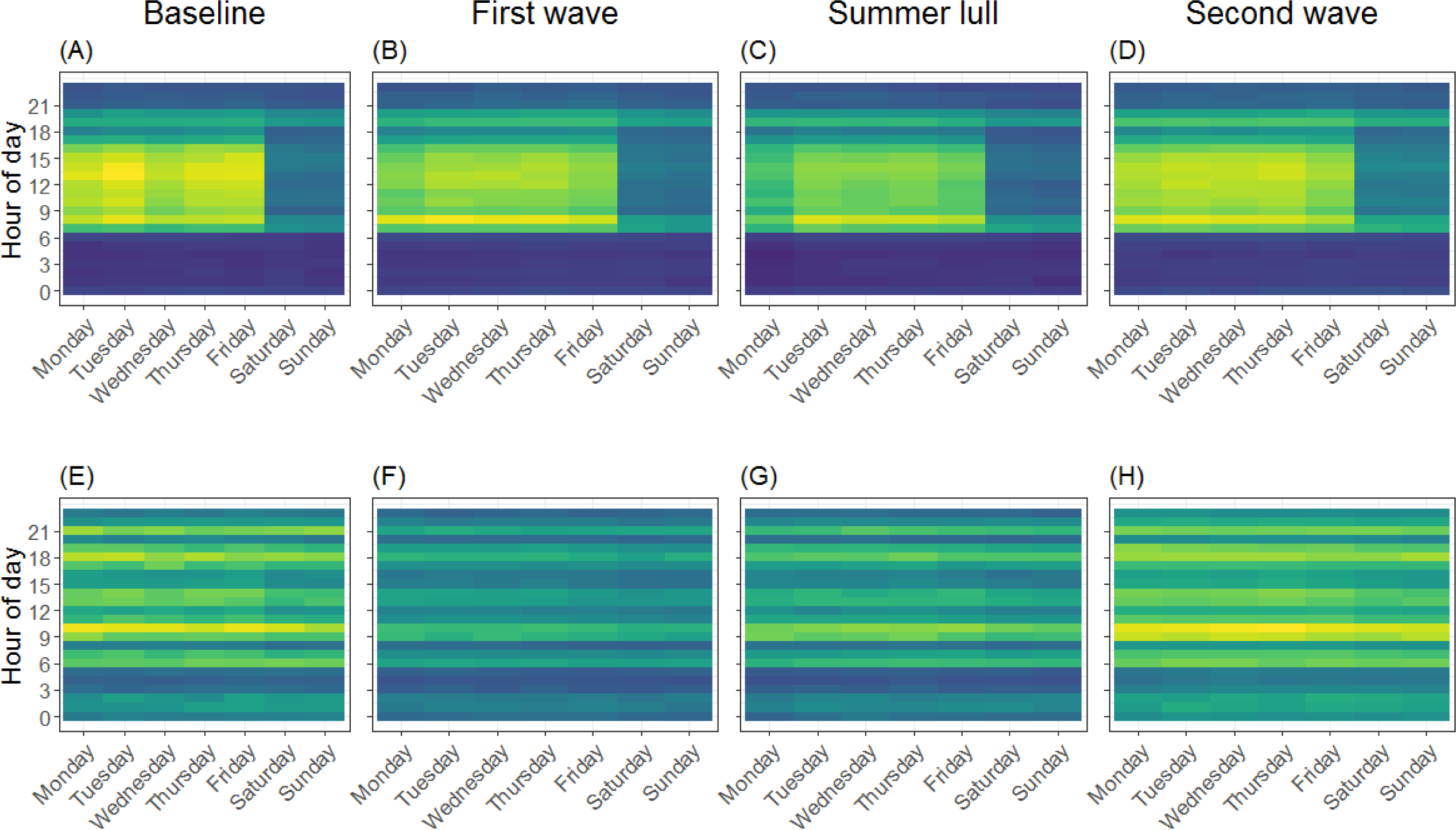
Heat maps for the number of door events and patient contacts at different hours of the day and week for healthcare workers at the Tower building of University College London Hospital during the COVID-19 pandemic. Plots A-D are for the number of door events and plots E-H are for patient contacts. Yellow cells represent a higher density of events and blue cells represent a lower density; to allow comparison between each stage of the pandemic, the colour gradient is relative to the maximum weight across all time periods.

During the first wave of COVID-19 patients (Figure 1C), the average daily number of staff with evidence of activity in the hospital fell by 10% compared to pre-pandemic levels. The average daily number of patients in the hospital dropped sharply, down by 44%. This coincided with a 24% decrease in the average daily number of patient contacts logged by HCWs, which was associated with a less prominent pattern in the weekly and hourly counts of contacts (Figure 2F). However, the per patient rate of daily contact was 41% higher than at baseline (i.e. on average patients in the first wave had more contact events with HCWs per day than that logged pre-pandemic). In contrast, the average daily number, rate and hourly pattern of door events remained relatively consistent 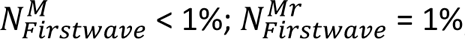; Figure 2B), which is surprising given the reduced staff levels.

In the summer lull, when there were fewer COVID-19 patients in the hospital, the average daily staffing levels and patient numbers remained below baseline levels 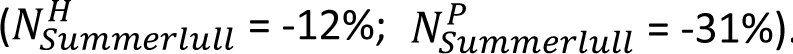. While the daily number and hourly pattern of patient contacts began to return towards that seen pre-pandemic, the average daily count of events remained lower than baseline levels 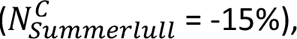, and the rate of contact was maintained above that seen at baseline 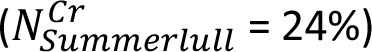. The average daily number of door events was 8% lower than baseline levels, and this was due to lower weekday peaks, while the rate of mobility remained similar to the pre-pandemic rate 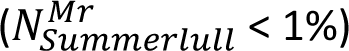.

During the second wave, both the average daily number of patients and staff in the hospital remained lower than that at baseline 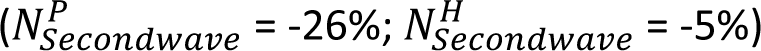. However, it is worth noting how daily staffing levels, after an initial drop during the Christmas break, followed the rise and fall of COVID-19 patients in the hospital, emphasising a different strategy by the hospital to that in the first wave where staff numbers were reduced. The second wave of COVID-19 patients also coincided with an increase in the daily number and rate of patient contacts and door events, all of which exceeded baseline levels 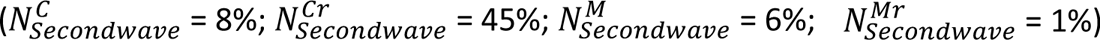

### Spatial-temporal dynamics

The pattern in the rate of mobility (as measured by the daily number of door events corrected for the number of HCWs) and rate of patient contacts (as measured by the daily number of logged patient contact events corrected for the number of patients) for HCWs on each floor of the Tower, differed in response to the pandemic (Table 3). Here we focus on the behaviour of HCWs on the six floors identified as key COVID-19 wards and that handled the majority (>= 15%) of COVID-19 patients (Figure 3, see Figure S1 for non COVID-19 wards); the AMU (floor 1), critical care (floor 3), HASU (floor 7), respiratory diseases (floor 8), general surgery (floor 9) and CoE (floor 10).

**Figure 3:**
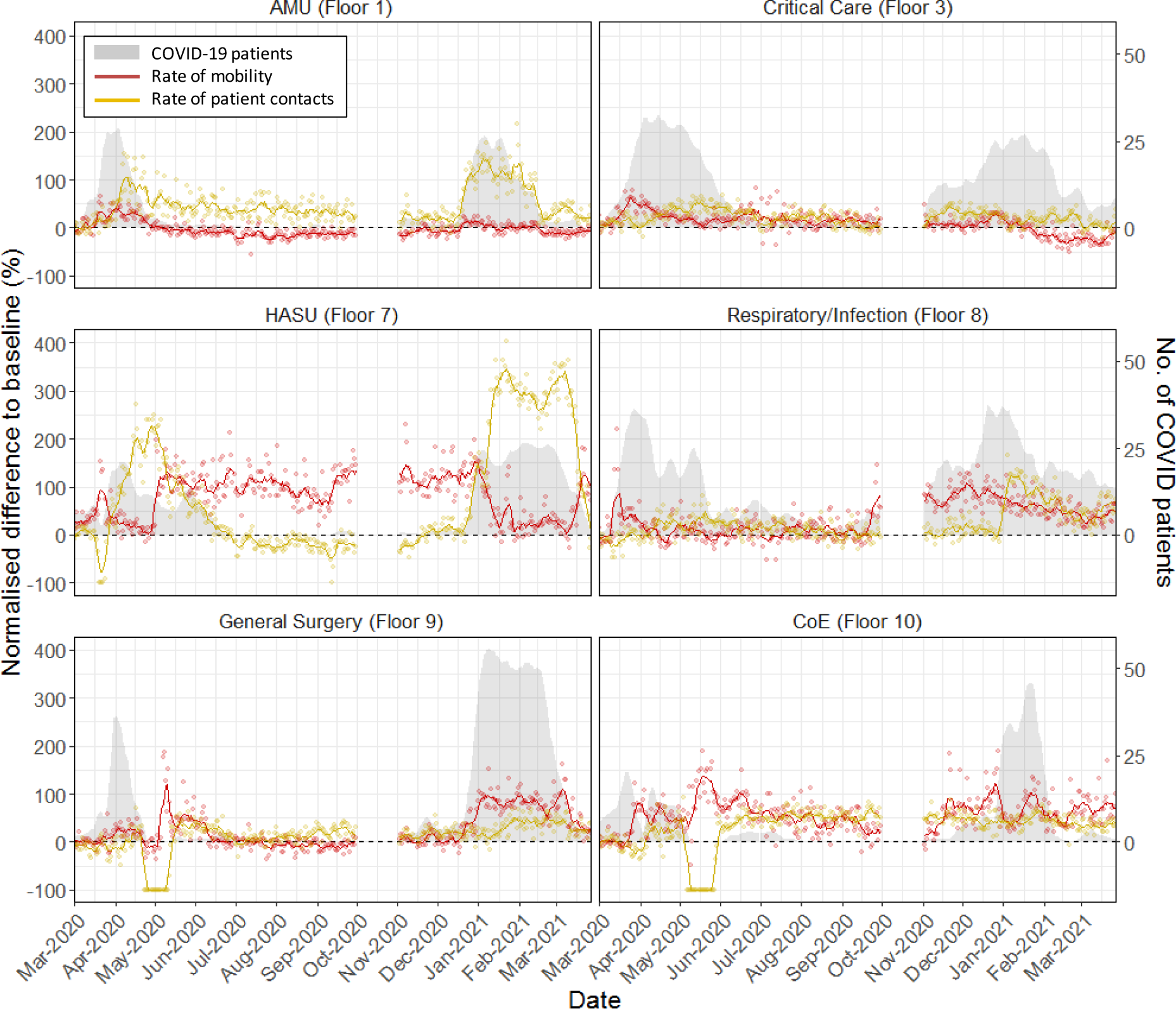
Changes in the rate of HCW mobility and patient contact on COVID-19 wards in the Tower building at University College London Hospital. The rate of healthcare worker mobility was measured in terms of the daily number of door events per healthcare worker, and the rate of patient contact was measured in terms of the daily number of logged patient contacts per patient. COVID-19 wards were identified as those that had >=15% of all COVID-19 patients (grey polygon) during the observation period. For each of the two measures, the normalized difference (N) to the average daily rates during the pre-pandemic (red/yellow points, representing percentage change) was calculated, and a smooth was applied using a seven day rolling average (red/yellow line). The black dotted line represents 0% change compared to the average in the pre-pandemic period. Data for October 2020 was not available.

**Table 3:** Changes in the rate of healthcare worker mobility and patient contacts in the Tower building at University College London Hospital during the COVID-19 pandemic. The rate of mobility (as measured by the average daily number of door events per healthcare worker), and the rate of patient contact (as measured by the average daily number of patient contacts per patient) during the pre-pandemic period (baseline) are presented for each floor of the Tower building. For each stage of the pandemic (First wave, Summer lull and Second wave) the normalized difference (N) to the average daily rates during the pre-pandemic stage are provided and expressed as percentage change. Rows in bold identify floors that handled the majority of COVID-19 patients (>=15%) during the pandemic.

| Floor | Mobility |  |  |  | Patient contacts |  |  |  |
| --- | --- | --- | --- | --- | --- | --- | --- | --- |
|  | Baseline | First wave | Summer lull | Second wave | Baseline | First wave | Summer lull | Second wave |
| Floor -1; Imaging | 2.4 | 7% | -3% | 10% | 6.4 | -29% | 7% | -11% |
| Floor G; ED | 9.4 | 18% | 2% | 8% | 12.7 | 20% | 30% | -68% |
| **Floor 1; AMU** | **5.8** | **7%** | **-15%** | **-3%** | **19.6** | **45%** | **35%** | **58%** |
| Floor 2; Day surgery | 3.1 | 17% | -6% | 1% | 18.8 | -4% | 30% | 44% |
| **Floor 3; Critical care** | **4.2** | **25%** | **15%** | **-4%** | **98.6** | **28%** | **16%** | **20%** |
| Floor 5; Nuclear medicine | 13.4 | -5% | -8% | -14% | 3.8 | -46% | -12% | 4% |
| Floor 6; Short stay surgery | 4.9 | 7% | 56% | -4% | 21.9 | 26% | 42% | 20% |
| **Floor 7; HASU** | **2.5** | **72%** | **101%** | **78%** | **34.6** | **73%** | **-21%** | **155%** |
| **Floor 8; Respiratory disease** | **7.1** | **14%** | **12%** | **64%** | **23.5** | **13%** | **7%** | **46%** |
| **Floor 9; General surgery** | **3.4** | **21%** | **-5%** | **50%** | **24.3** | **-7%** | **13%** | **26%** |
| **Floor 10; CoE** | **3.1** | **59%** | **43%** | **66%** | **24.8** | **-2%** | **53%** | **44%** |
| Floor 11; Paediatrics | 7.6 | 17% | 13% | 25% | 14.2 | 33% | 26% | 28% |
| Floor 12; Adolescents | 4.5 | 52% | 16% | -1% | 19.7 | -30% | 34% | 44% |
| Floor 13; Oncology | 3.3 | 11% | 22% | 55% | 22.1 | 24% | 28% | 28% |
| Floor 14; Head and neck | 4.7 | 0% | 8% | -8% | 23.3 | 21% | 31% | 27% |
| Floor 15; Private wards | 9.3 | -14% | -27% | -22% | 9.4 | -41% | -31% | 51% |
| Floor 16; Haematology | 7 | -7% | -1% | -2% | 30.5 | 12% | 16% | 11% |

The daily rate of mobility for HCWs during the first wave was, on average, higher on all COVID-19 floors compared to baseline levels. During the summer lull, the daily average rate of mobility on AMU and General surgery fell below baseline levels, while HCW mobility on all other COVID-19 floors increased, with the most notable increase on HASU 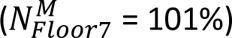. In response to the second wave of COVID-19 patients, the rate of mobility increased further above baseline levels on floors with HASU, Respiratory disease, General surgery and CoE, while HCW mobility was only marginally below baseline levels on AMU and Critical care.

During the first wave and compared to pre-pandemic levels, the average daily rate of patient contact increased on AMU, Critical care, HASU and Respiratory disease. In contrast, the rate of patient contact was lower on average for General surgery and CoE, both of which had a period of days in May with zero contacts logged, which may indicate that the floors were closed during this period. In the summer lull and with the exception of HASU, the average daily rate of patient contact was higher than baseline levels for all COVID-19 floors. The rise in COVID-19 patients in the second wave coincided with a further increase in the rate of patient contact on all COVID-19 floors, with the most notable increase on HASU 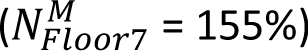.

### Spatial connectivity

The connectivity between floors (based on the number of HCWs that had activity on any two floors in the same day) revealed that some were more connected than others, and that the resulting clustering of floors varied throughout the pandemic (Figure 4). During the baseline period, three clusters were identified; one large cluster containing Imaging through to Plant (floors -1 to 4), Short stay surgery, HASU and General surgery; a smaller cluster comprising the Basement, Nuclear medicine, Paediatrics and Adolescents; and a third consisting of Respiratory disease, CoE and Oncology through to Haematology (floors 13 to 16).

**Figure 4:**
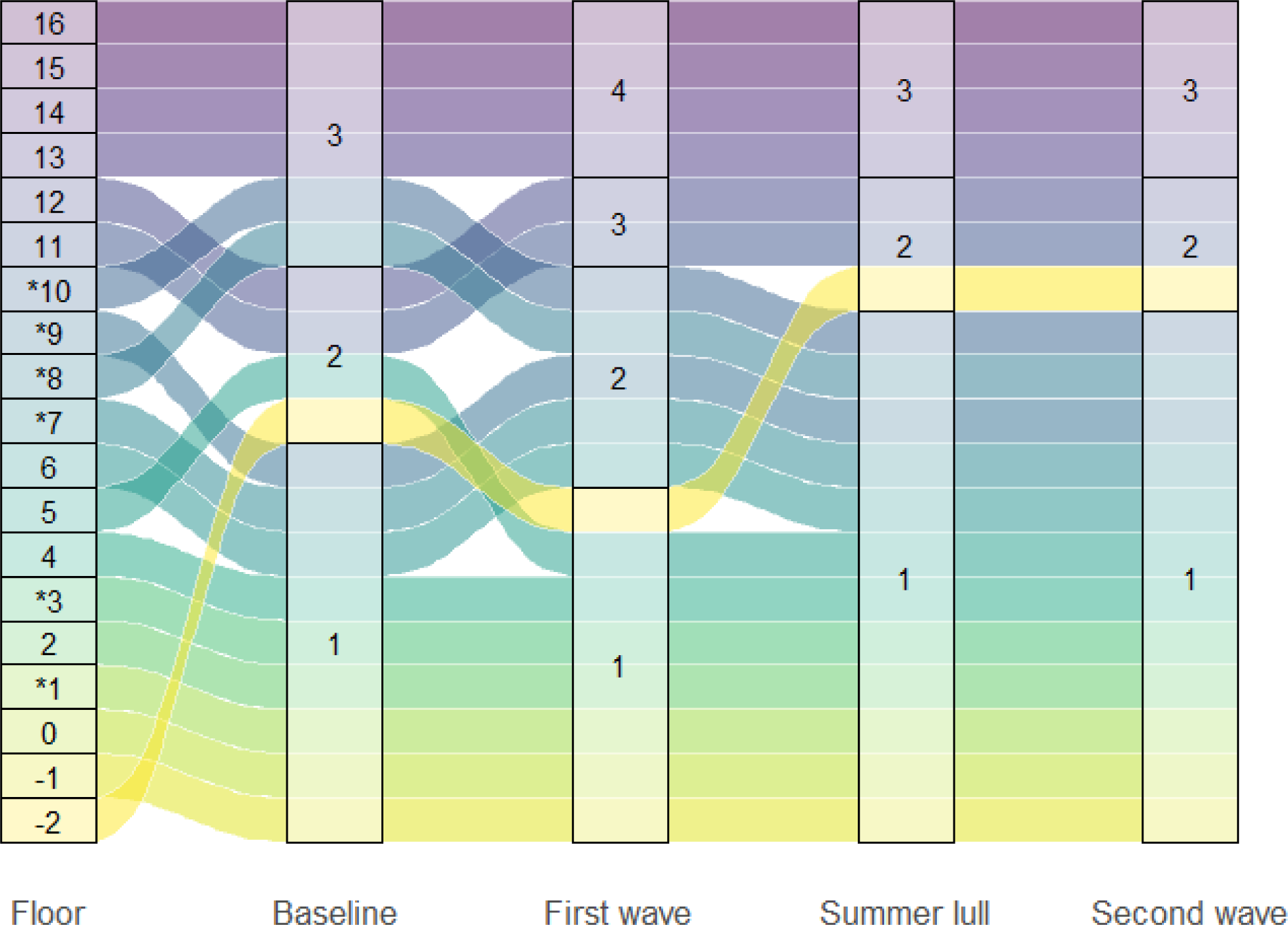
The spatial connectivity of floors in the Tower building at University College London Hospital during the COVID-19 pandemic. The alluvial diagram depicts the connectivity of floors, as determined from the Louvain clustering algorithm and using dyadic weights derived from the number of healthcare workers with logged door events and/or patient contacts on the focal floors during the same day. The numbering in the left most column identifies the floors, and the numbers in the remaining four columns represent the cluster group the floors belong to in each stage of the pandemic; pre-pandemic (baseline), first wave, summer lull and second wave. An asterisk identifies floors that had the majority (>=15%) of COVID-19 patients during the observation period.

During the first wave, the connectivity between floors changed such that four clusters were identifiable, and floors adjacent to each other were generally in the same cluster. The basement through to Nuclear medicine (floors -2 to 5) formed a cluster, while Short stay surgery through to CoE (floors 6 to 10) formed a second cluster, Paediatrics and adolescents (floors 11 and 12) made up a third cluster and the forth consisted of Oncology through to Haematology (floors 13 to 16). Floors identified as COVID-19 wards were present in two of the four clusters, which also included non COVID-19 wards.

Adjacent floors were again more likely to be connected during the summer lull, but only three clusters were identified; Imaging through to CoE (floors -1 to 10) formed the largest cluster, while the basement, Paediatrics and Adolescents (floors -2, 11 and 12) were in a second cluster, and the third cluster consisted of Oncology through to Haematology (floors 13 to 16). During the second wave the connectivity of floors and the clusters they formed were unchanged from that in the summer lull, suggesting that the spatial activity of HCWs had stabilised. All COVID-19 floors were in the larger cluster, which also included non COVID-19 wards.

### Indirect contacts between patients

The average daily patient connectivity (due to shared contacts with HCWs on the same day), showed daily fluctuations that were not consistent during the course of the pandemic (Figure 5). The average daily connectivity of COVID-19 negative patients with other COVID-19 negative patients 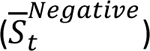 remained stable throughout the pandemic 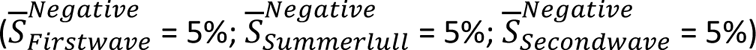 however, during the first wave the pattern in *S*_*t*_^*Negative*^ followed the rise and fall of COVID-19 patients in hospital before settling around 5% in the summer lull where it remained until the end of the observation period.

**Figure 5:**
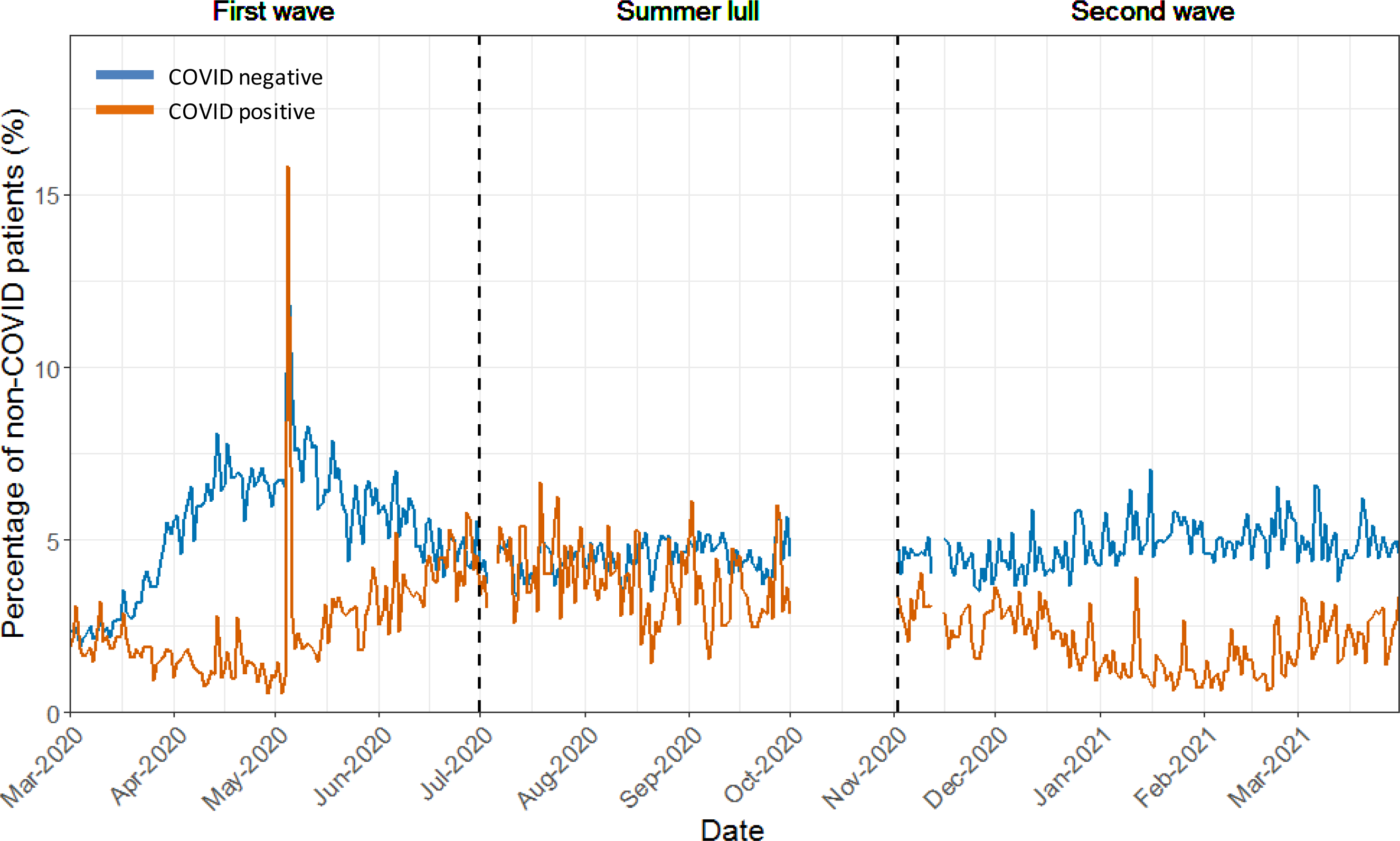
The average daily percentage of COVID-19 negative patients that had indirect contacts with other patients in the Tower building at University College London Hospital during the COVID-19 pandemic. The daily averages are plotted separately for the percentage of COVID-19 negative patients to have indirect contacts with COVID-19 positive (orange) and other COVID-19 negative (blue) patients. An indirect contact was determined through evidence of a shared contact with HCWs on the same day. Data for October 2020 was not available.

In contrast, during the first wave, the hospital reduced the daily connectivity between COVID-19 negative patients and COVID-19 positive patients (*S*_*t*_^*Positive*^) to a low of <1%. However, 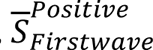 was 2%, and there was a noteworthy spike in *S*_*t*_^*Positive*^ to 16% on the 5^th^ May 2020, which was due to one HCW who had 89 patient contacts; 33 of which were positive for COVID-19. During the summer lull, much greater levels of indirect contact between patient groups was seen 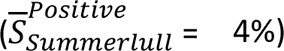, suggesting a relaxation in staff cohorting procedures. During the second wave, *S*_*t*_^*Positive*^ gradually dropped to a low of <1% and 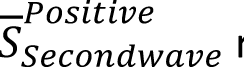 returned to 2%, highlighting a reactive response to the rise of COVID-19 patients.

## Discussion

Mobility and contact rates are fundamental to the transmission of communicable diseases, and data on these behaviours are extremely valuable for epidemiological investigations. It has long been established that HCWs can be part of transmission clusters within healthcare settings (Huttunen & Syrjänen, 2014; Lumley et al., 2021) however, data on the behaviour of HCWs are often scarce. In this paper, we demonstrate how behavioural markers for HCW mobility and patient contacts within the hospital, can be derived from EMRs and door access logs at an aggregate level. Using data from a London teaching hospital and during the COVID-19 pandemic, we provide a framework to further support IPC practitioners in assessing patterns of staff behaviour, identifying behavioural change and in conducting evidence-based infection control.

The temporal trends in workforce and HCW behaviour are in line with those reported in other studies (Champredon et al., 2018; Duval et al., 2018; Gallego et al., 2015; Vanhems et al., 2013), and illustrate the utility of the featured data sources in representing the working patterns of HCWs. Staff and patient levels determined daily patterns in HCW behaviour, and this was evident when the hospital reduced staff and patient numbers during the first wave of COVID-19 patients, which resulted in a notable drop in logged patient contacts. However, the rate of patient contact (number of contact events per patient) was maintained above baseline levels throughout the pandemic, while the rate of HCW mobility (number of door events per HCW) remained relatively stable, only surpassing baseline levels in the second wave. Causes for the observed changes in HCW behaviour are hard to ascertain, but are likely products of shifting working practices (e.g. through IPC activities), perceptions of risk (e.g. before/after vaccination and changes in the availability of PPE) and hospital pressures (e.g. needs of the patient population). Even without clear causal pathways, it is evident that these data can be used to monitor the behaviour and activity patterns of the HCW population, providing novel insights for IPC practitioners.

Patterns of HCW behaviour showed significant spatial variation in response to the pandemic. Increases in the rate of mobility and rate of patient contact were most notable on COVID-19 floors during the first and second waves. However, the degree of change in these behavioural markers was not equal across floors and, despite few (or no) COVID-19 patients, non COVID-19 floors also experienced changes in HCW behaviour. Trends in HCW behaviour on different floors will depend on the functions of the wards occupying them, how these functions evolved during the course of the pandemic and on IPC interventions. Combining observations of how HCW behaviour varied spatially with data on the context in which they occurred, will be necessary to help explain spatial heterogeneity in the infection risk for HCWs, as was seen during the pandemic (Gómez-Ochoa et al., 2021).

One strategy to prevent nosocomial transmission is to cohort patients and staff, whereby patients positive for the disease of concern and/or the staff responsible for their care, are kept separate to the rest of the patient population (Ahmad & Osei, 2021). At UCLH this was achieved by establishing COVID-19 wards that handled the majority of COVID-19 patients. Using the routinely-collected data we were able to identify the main COVID-19 wards and monitor the daily indirect contacts between patients (as determined through shared contacts with HCWs on the same day). Successful staff cohorting would have resulted in no indirect contact between COVID-19 negative and positive patients. However, this was not consistently achieved and, while the indirect contacts between these groups of patients were substantially reduced during the first and second waves, the response was not maintained during the summer lull, and appears reactive to increases in the number of COVID-19 patients. Staff cohorting can be prevented by numerous practical limitations, and the pandemic presented many challenges including staff shortages. That said, the routinely collected data provides a tool for IPC practitioners to monitor the success of interventions such as cohorting, and offers a means to quickly identify, investigate and react to undesirable spikes in indirect patient contacts that could compromise patient and staff safety.

Another strategy available to IPC practitioners is to limit the traffic within the hospital (Ahmad & Osei, 2021). At UCLH a number of interventions were adopted to accomplish this, including reduced patient and staff numbers, and through the installation of ‘COVID doors’ that created barriers to disrupt the flow of people between spaces. Our analyses identified the reduced staff and patient numbers, but we were unable to assess the effect of COVID doors on the mobility of HCWs, as the dates of their installation and use were not known. Instead, we assessed the flow of HCWs between floors and found that, after some fluctuation in the early stages of the pandemic, the connectivity between floors stabilised, suggesting a new ‘normal’ to the working practices of HCWs. Throughout the pandemic, the flow of HCWs continued between COVID-19 and non COVID-19 wards, which may highlight an opportunity to improve IPC activities. However, a higher resolution analysis that takes into account the partitions within floors may reveal the true flow of staff between COVID-19 and non-COVID-19 areas.

In this investigation we used a minimal number of data fields and metrics aggregated at the level of the HCW population. However, further insights into the variations of HCW behaviour could be uncovered if the data were paired with other data fields and aggregated by individual or HCW group. For example, combining this data with specific details on where and when IPC interventions were introduced, would allow investigations into the pre and post effects of interventions on HCW mobility and patient contacts. Combining these data sources with data on the disease status of staff could help identify HCW groups and/or individuals more at risk of acquiring HAIs, along with the behaviours or working conditions associated with higher risk. As healthcare systems move towards more digital systems, the accessibility and diversity of data available to practitioners grows, providing new opportunities for research and support for evidence based infection control.

While the data sources featured here have potential to be used operationally by IPC practitioners, there are several challenges that hospitals may have to overcome for this to be realised. Firstly, it is worth noting that UCLH is a digital hospital, but many healthcare facilities in the UK and across the world are not, and an absence of electronic records will reduce the scope and capacity of facilities to utilise these data. Hospitals, particularly those within the NHS, often outsource services such as systems for security door logs and EMRs, and in this study the various datasets from outsourced companies had to be consolidated, which required the creation of a master staff index to establish links between the databases. Mapping the data flows and creating a user friendly platform will be challenging and requires the collaboration of researchers, IT professionals and IPC staff. There are also challenges in relation to the generation of these data themselves, as we lack assurances on the exact nature of the processes underlying their generation. For instance, the use of staff cards to open security doors may be biased in time and space by HCWs following each other through doors (e.g. during ward rounds), or by doors being left open. Likewise, there has been little systematic analysis to date on the generation of EMR data, in relation to the accuracy of the spatial or temporal markers, or the HCWs involved in events. These remain important validation challenges that are being undertaken – but nevertheless, the principles underlying aggregate patterns produced using these data appear sound.

Data on behaviours with epidemiological relevance are often scarce but, as hospitals embrace the digital age, data is becoming more readily available. Deriving behavioural markers from routinely collected data provides opportunities to enhance IPC activities aimed at better protecting HCWs and patients, in addition to improving pandemic preparedness. IPC practitioners should consider what new data sources are at their disposal and how they can be used operationally to empower decision making.

## Data Availability

Data will be made available upon request or via a data repository service after confirmation of acceptance in a peer reviewed journal.

## Acknowledgements

This study was supported by the UCLH/UCL NIHR Biomedical Research Centre and funding from the UKRI MRC (grant ref: MC_PC_19082), and UCLH Charity. For their support we thank the UCLH medical directors Charles House and Gill Gaskin, Pushpsen Joshi at the Joint Research Office, Nathan Lea from the UCLH information governance department, Leila Hail from the UCLH infection control department, Richard Clarke, David Ramlakhan, David Thompson and Gareth Adams at the UCLH digital services department, and all involved with the SAFER research programme.

## Author contributions

EN, MS, CH and EM supervised the project. EN and EM designed the research. WW, CL and SK extracted the data. JWA, EM and NG analysed the data. JWA and EM wrote the manuscript and all authors approved the final version.

## Data availability

Data will be made available via a data repository service after confirmation of acceptance.

## Ethics statement

The study protocol was approved by the NHS Health Research Authority (South Central – Berkshire REC ref 20/SC/0147, protocol number 130861) and ethical oversight was provided by the UCLH research ethics committee (IRAS project ID: ref. 281836). UCL GOS ICH R&D approval number 20PL06.

