## Supplementary material for "Using routinely collected hospital data to investigate healthcare worker mobility and patient contacts within a UK hospital during the COVID-19 pandemic"

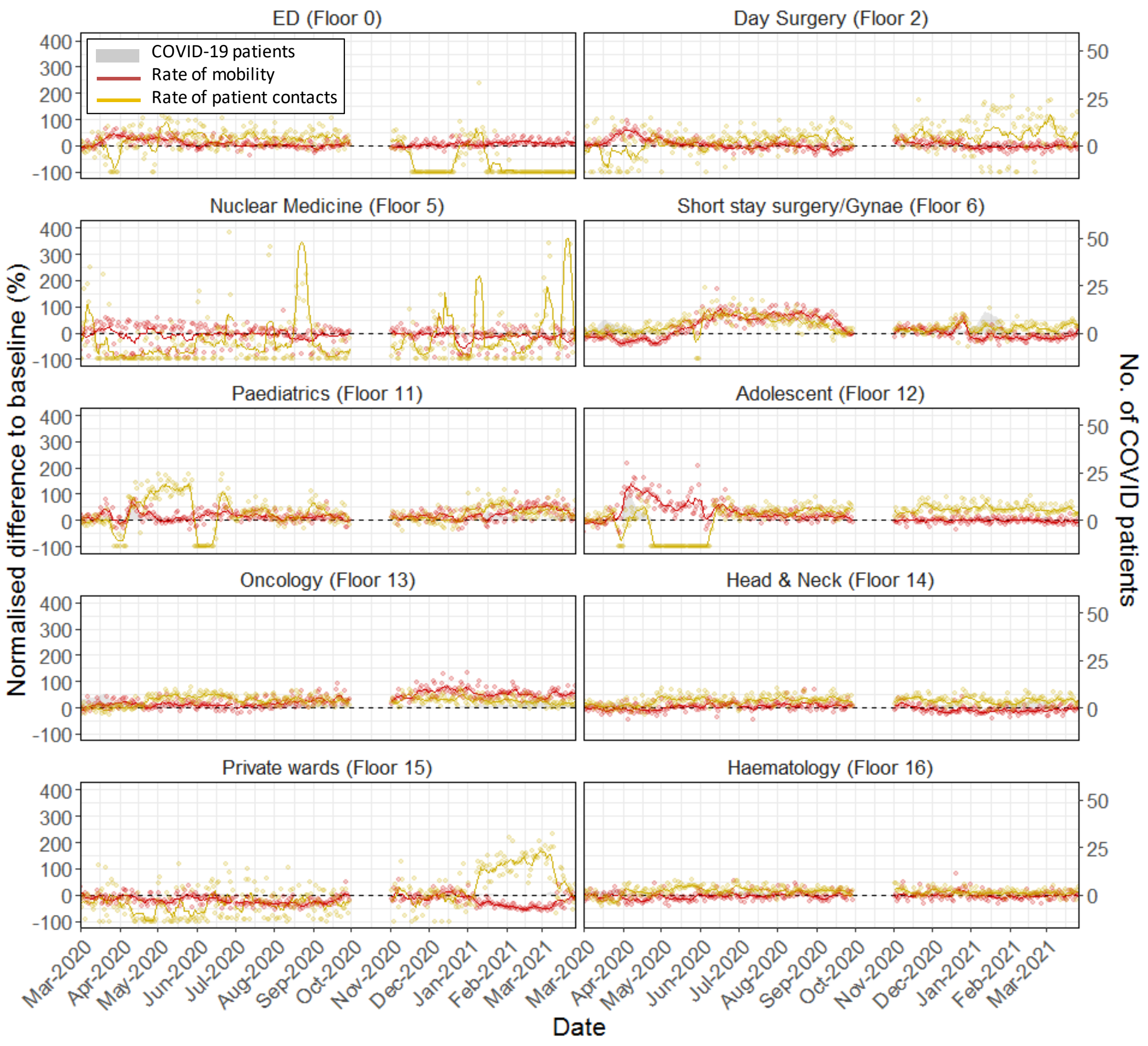


Figure S1. Changes in the rate of healthcare worker mobility and patient contact on non COVID-19 wards in the Tower building at University College London Hospital. The rate of healthcare worker mobility was measured in terms of the daily number of door events per healthcare worker, and the rate of patient contact was measured in terms of the daily number of logged patient contacts per patient. Non COVID-19 wards were identified as those that had <=15% of all COVID-19 patients (grey polygon) during the observation period. For each of the two measures, the normalized difference (*N)* to the average daily rates during the pre-pandemic (red/yellow points, representing percentage change) was calculated, and a smooth was applied using a seven day rolling average (red/yellow line). The black dotted line represents 0% change compared to the average in the pre-pandemic period. Data for October 2020 was not available.
